# Beyond the Door: Mapping Patient Experiences in an Open-door Inpatient Psychiatric Unit

**DOI:** 10.1101/2025.10.01.25337052

**Authors:** Jorge Cuevas-Esteban, Esther Ruiz-Garros, Encarnacion Romero-Diaz, Nicole Motta-Rojas, Beltran Jimenez-Fernandez, Aurea Fernandez-Ribas, Barbara Martinez-Cirera, Anna Moreno-Orea, Laura Miro-Mezquita

## Abstract

Open-door policies in inpatient psychiatric care have been increasingly adopted to enhance patient autonomy and reduce coercive practices. However, systematic methodologies for evaluating patient experiences in these settings remain limited. This study applies Patient Journey Mapping within an open-door acute psychiatric unit, to comprehensively analyze the hospitalization experience. Using a mixed-methods approach, qualitative data were first collected via focus groups (n=18), adhering GRAMMS guidelines, to identify critical touchpoints, and then quantitative data (n=32) were gathered through an adapted Likert-type ordinal scale to assess patient satisfaction. Data were analyzed using content analysis methodology. Results delineate key hospitalization phases and identify “Moments of Truth,” “Moments of Pain,” and “Wow Moments,” which significantly shape patients’ perceptions of care quality. While open-door policies promote autonomy and reduce psychological distress, challenges persist in admission processes, medication administration privacy, and communication regarding structured activities. These findings highlight the need for patient-centered methodologies in psychiatric care to optimize service delivery, foster therapeutic environments, and enhance recovery-oriented practices.

## 1. Introduction

In recent years, mental healthcare has undergone a paradigm shift toward patient-centered models, prioritizing the reduction of coercive interventions and the promotion of recovery-focused care (Tingleff et al., 2017). Traditional closed-door policies in psychiatric inpatient units, historically justified as safety measures (Van Der Merwe et al., 2009), have been linked to critical issues such as increased patient distress (e.g., perceptions of powerlessness and stigmatization)(Bakolis, Gupta and Wykes, 2022; Butterworth, Wood and Rowe, 2022), restricted autonomy (e.g., limited participation in care decisions) (Steinauer et al., 2020; Bakolis, Gupta and Wykes, 2022), and lower satisfaction with treatment(Bowers et al., 2010). For instance, locked environments in dual-diagnosis units correlate with higher patient agitation and reduced therapeutic engagement (Steinauer et al., 2020), while surveys reveal that both patients and staff associate closed-door practices with heightened tension and mistrust (Bowers et al., 2010; Hotzy et al., 2018).

Conversely, open-door policies in psychiatric settings have shown to promote therapeutic alliances, decrease incidents of seclusion and forced medication (Jungfer et al., 2014; Hochstrasser et al., 2018), and enhance patient-reported outcomes, such as perceived dignity and collaboration in care (Indregard et al., 2024). Empirical evidence from randomized trials, such as the Norwegian study by Indregard et al.(Indregard et al., 2024), demonstrates that open-door approaches maintain safety standards while reducing coercive measures. Similarly, longitudinal implementations of these policies report sustained reductions in seclusion rates over six years (Hochstrasser et al., 2018). However, the implementation of open-door psychiatric units presents unique challenges, particularly in balancing safety with the promotion of patient autonomy and dignity (J. Kalagi et al., 2018). Patients generally view open-door policies positively, as they offer a sense of autonomy and reduce feelings of stigmatization (Schreiber et al., 2024). The open-door concept is often seen as a therapeutic tool that promotes patient-centered care. Patients appreciate the opportunity to engage in meaningful activities and interact with peers, which can foster a sense of community and support (J Kalagi et al., 2018; Cooper et al., 2023).

Patient Journey Mapping has emerged as a methodology that helps healthcare professionals deeply understand the patient experience by visually mapping their path through the healthcare system. By identifying critical touchpoints, both positive and negative, throughout the care process, this mapping technique enables healthcare providers to design interventions that enhance patient engagement, improve service delivery, and support organizational change (Bulto et al., 2024; Guilcher et al., 2024). This approach integrates qualitative and quantitative data to highlight pain points, uncover opportunities for improvement, and facilitate the co-design of patient-centered interventions (Gualandi et al., 2019; Bulto et al., 2024). As highlighted in recent work by Sijm-Eeken et al., there is currently no universally accepted methodology for conducting patient journey mapping, and substantial heterogeneity exists in the rigor and quality of the approaches employed across studies (Sijm-Eeken, Zheng and Peute, 2020). While the mapping of patient journeys facilitates the identification of barriers, enablers, and service delivery gaps from a patient-centred perspective, major shortcomings in the reporting of these initiatives continue to limit both transparency and replicability(Davies et al., 2023).

To the best of our knowledge, this study represents the first application of Patient Journey Mapping within an open-door psychiatric hospitalization unit. By employing a mixed-methods approach to capture patient perspectives, this research aims to provide actionable insights for improving patient experiences and fostering a more therapeutic care environment. Specifically, the objectives of this study are to: define the key stages of the patient journey within an open-door psychiatric unit, document patient perceptions using a structured rating scale, identify non-value-adding and dispensable actions to optimize unit dynamics, propose targeted improvement actions based on patient experiences, and visually represent the patient journey to facilitate comprehension and dissemination of findings.

## 2. Methods

### 2.1. Setting

This study was conducted in 2022 at the open-door inpatient psychiatric unit of Germans Trias University Hospital (GTUH) in Badalona, Barcelona, Spain. This facility is a general multispecialty hospital affiliated with the Autonomous University of Barcelona. At the time of the study, the GTUH served a catchment area encompassing 350,530 adults, distributed among four community mental health zones within the North Barcelona health district. At the time of the study, the acute inpatient psychiatric unit of the hospital functioned as a single ward with an operational capacity of 14 beds, staffed by a multidisciplinary team that includes senior and junior psychiatrists, nurses, psychologists, and social workers. The unit delivers comprehensive acute care for individuals with severe mental illnesses, utilizing a combination of pharmacotherapy, psychotherapy, social work, occupational therapy, and recovery-focused programs. Since 2019, the unit has progressively implemented the Safewards care model. Safewards is a set of ten interventions designed to enhance safety by preventing conflict and containment (Bowers et al., 2015). Electroconvulsive therapy was available at the time of the study. Most patients (90-95%) are admitted from the emergency room. Patients typically present with severe primary psychiatric disorders characterized by acute disturbances and/or a risk of self-harm. The remaining individuals are referred by community mental health services. Of the participants included in the study, 60 % were admitted involuntarily under applicable mental health legislation, while 40% were voluntary admissions.

At the time of the study, the unit operated with an approximate staff-to-patient ratio of 1:7 during daytime shifts and 1:10 during night shifts. The unit also implemented an open-door policy that permitted patients to move freely in and out totaling approximately 6 hours of open access per day. This schedule was consistently maintained throughout the week, with no major deviations during the study period. This approach has been in place since October 2021.

### 2.2. Design

This study employed a mixed-methods approach to comprehensively map the patient’s journey in an open-door psychiatric hospitalization unit with the Safewards care program. The design integrated qualitative data collection and analysis to identify strengths and weaknesses at each hospitalization stage, complemented by a quantitative study to assess patient satisfaction with the moments and the staff. This approach provided a comprehensive understanding of the patient’s trajectory from initial contact with the mental health unit through to discharge. After analyzing all collected data, a patient journey map (PJM) was developed from the perspective of the patients.

Before conducting this mixed study, a multidisciplinary team of mental health hospitalization experts, including one psychiatrist, two psychiatry residents, one social worker, one psychologist, one nurse, and one auxiliary nurse, defined the starting, middle stage, and ending stages and moments that constitute the patient’s experience during mental health hospitalization in the open-door unit. They identified fifteen distinct moments grouped into five stages.

#### 2.2.1. Qualitative phase

To analyze patients’ perspectives, two focus groups with patients (total n=18) from the unit were conducted, each one divided into two sessions lasting approximately one hour (total of 4 hours).The number of participants was determined by the total number of patients available and willing to participate at the time of the study. The participants in the same focus group sessions remained largely consistent, with minor variations due to scheduling conflicts, and were held at the hospital facilities. The focus groups were integrated into the unit’s regular group routines; however, participants were explicitly informed that participation in the research was separate and distinct from their usual therapeutic activities. Information about participants and the duration of each session can be found in Table 1. The inclusion criteria for participants were as follows: be a patient admitted to the hospital’s acute mental health unit, be over 18 years old, and basic proficiency in Spanish or Catalan to understand the interview questions. A total of 28 patients hospitalized in the unit during the study period were approached in person and invited to participate in the study; however, 10 declined due to their psychopathological conditions. Verbal informed consent were obtained from all participants after they were provided with detailed information about the study’s objectives, procedures, confidentiality measures, and their right to withdraw at any time without affecting their care. Participation remained entirely voluntary. The study adhered to ethical guidelines, receiving approval from the local research ethics committee (PI-25-133).

**Table 1.** Duration, Number of participants, Sex (% female), Average age (SD) of the participants of the focus groups.

|  | **Duration** | **Number of participants** | **Sex (% female)** | **Average age (SD)** |
| --- | --- | --- | --- | --- |
| Group 1 | 60 minutes | 10 | 60% | 40.4 |
|  | 45 minutes | 8 | 40% | 44 |
| Group 2 | 60 minutes | 8 | 25% | 56.5 |
|  | 60 minutes | 8 | 40% | 52 |

The primary objective was to deepen the understanding of how the open-door mental health unit operates, with a focus on identifying elements that enhance and detract from the patient experience, as well as determining which hospital practices are effective and which require improvement. This approach enabled the development of the experience map section, which details both the strengths and critical areas of the hospitalization process from the patient’s perspective. The interviews were conducted by a psychiatrist and a social worker from the multidisciplinary team, with the social worker serving as the note-taker. No other individuals were present in the interview room except for the participants and the interviewers. Interview guides were developed based on a literature review and refined after initial interviews to ensure relevance and capture detailed experiences. All participants provided written informed consent prior to participation, and measures were taken to ensure anonymity and confidentiality. To effectively capture and visualize the patient’s journey, the dynamics of the sessions were centered around answering the questions, “What does the hospital do well?” and “What does the hospital do poorly?” for each moment identified by the expert panel. Visual tools were used to help clarify the questions and guide participants’ responses at each stage. Moments and stages previously defined by the expert panel were refined during the focus groups according to patient’s opinions. Direct quotes from the participants were collected to enrich the data.

#### 2.2.2. Quantitative phase

After completing the qualitative phase of the study in March 2022, the research progressed to the quantitative phase. To evaluate patients’ subjective experience across key touchpoints of the care process, a structured instrument was developed by the research team based on an adapted Likert-type ordinal scale. This scale was designed not only to measure satisfaction levels but also to capture the emotional intensity associated with patient perceptions at each stage of the healthcare journey. The traditional Likert scale, originally developed by Rensis Likert in 1932, has been widely used in social and health sciences to measure attitudes and opinions through ordered response categories (Likert, 1932). In this study, a 6-point scale ranging from highly negative to highly positive emotional descriptors (e.g., “I hate it” to “WOW”) was employed, which provides a more intuitive and emotionally resonant response format. This approach draws from principles of the semantic differential technique developed by Osgood et al. (Osgood et al. 1957), where evaluative language is used to elicit connotative meaning. Such emotionally enriched adaptations have been increasingly applied in patient-centered research, particularly in-service design and user experience (UX) evaluations, where traditional Likert scales may lack the expressive granularity required to assess affective responses. Emotional labeling of response options can improve engagement and response accuracy, especially when mapping complex, subjective experiences such as vulnerability, empathy, or perceived professionalism in healthcare interactions (Boone and Boone, 2012).

The scales focused on patient satisfaction regarding “Experience in Interactions” and “Empathy and Professionalism” at each stage and moment of hospitalization, with participants rating each moment on a Likert scale from 1 to 6, where 1 indicated the worst experience and 6 indicated the best. These scales, which can be found in Appendix I, were completed by patients prior to hospital discharge at the hospital facilities. The general inclusion criteria were identical to those of the qualitative phase, except that participants who had taken part in the focus groups were excluded. All eligible participants received detailed face-to-face information about the study’s purpose and scope during a preliminary session, and informed consent was obtained from all subjects. A total of 32 assessments were conducted by a psychiatrist and two psychiatry residents, with an average duration of 20 minutes per assessment. The mean participant age was 25 years, and 60% were female. The sample size was determined pragmatically based on the number of eligible and consenting patients during the study period, in line with the exploratory and complementary nature of this phase within the mixed-methods design.

### 2.3. Data Analysis

The qualitative data were analyzed using a rigorous content analysis approach to uncover aspects of care that enhance or detract from the overall patient experience across the various stages of hospitalization. Focus group information was independently coded by two experts, capturing both the strengths and the areas needing improvement in patient care. These initial codes were reviewed and refined in a series of iterative discussions, during which similar codes were grouped per hospitalization stage into themes. The emerging themes were subsequently validated through member checking. The study adhered to the Good Reporting of A Mixed Methods Study (GRAMMS) guidelines, ensuring transparency and methodological rigor (O’Cathain, Murphy and Nicholl, 2008).

During the analysis, the team identified critical junctures in the patient journey, which were classified into three key categories: “Moments of Truth,” “Moments of Pain,” and “Wow Moments.” Moments of Truth refer to highly impactful interactions where patients critically evaluate hospital services, influencing both their satisfaction and adherence to treatment. In contrast, Moments of Pain emerge from service delivery failures or negative perceptions that detract from the overall patient experience. Finally, Wow Moments are exceptional experiences that provide significant added value and often lead to positive word-of-mouth. Table 2 presents the definitions of these moment classifications and the corresponding symbols used in the PJM.

**Table 2.**
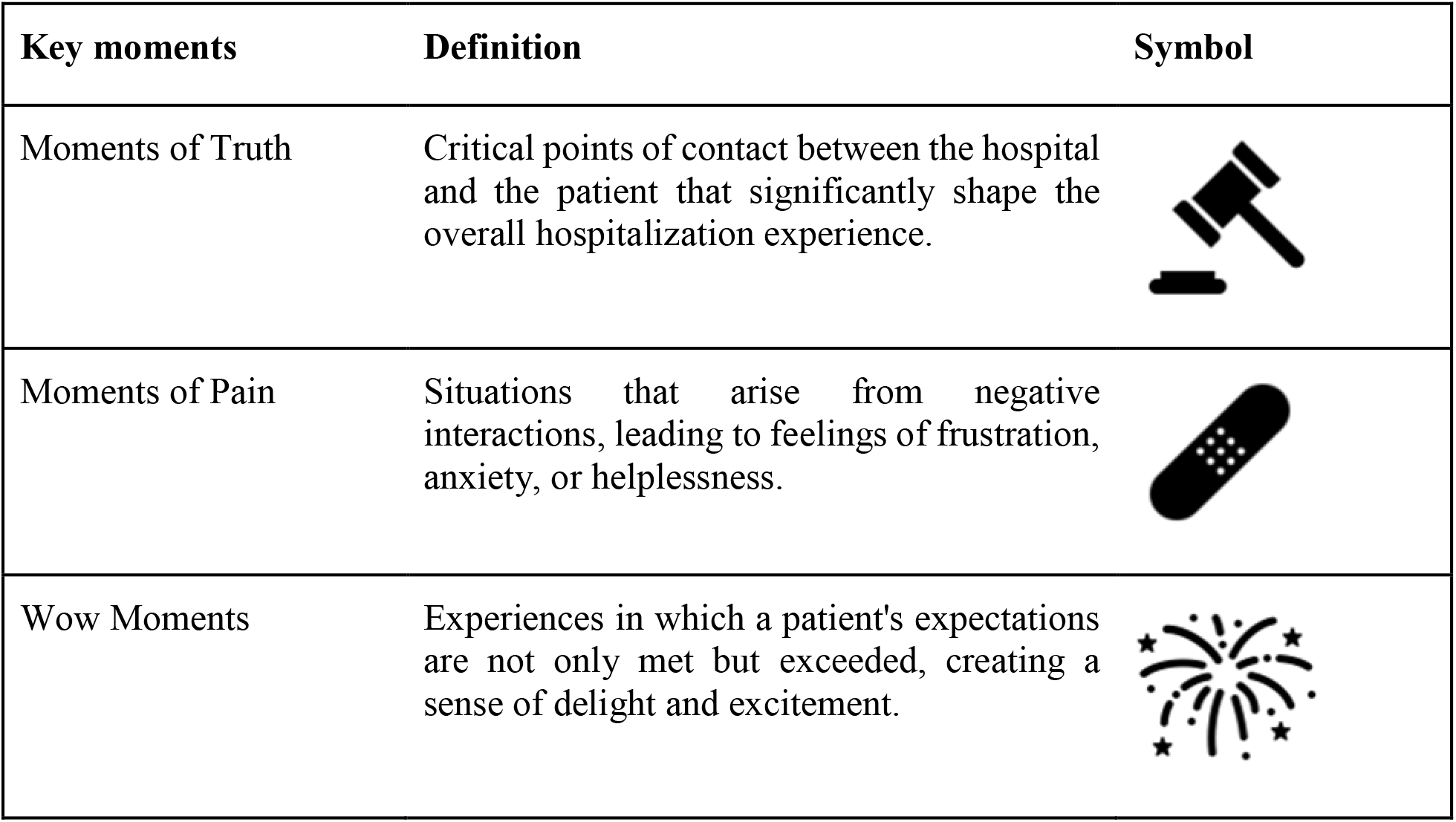
Definition of Key Moments of the patient experience during hospitalization and their symbolic representation in the PJM.

| Key moments | Definition | Symbol |
| --- | --- | --- |
| Moments of Truth | Critical points of contact between the hospital and the patient that significantly shape the overall hospitalization experience. |  |
| Moments of Pain | Situations that arise from negative interactions, leading to feelings of frustration, anxiety, or helplessness. |  |
| Wow Moments | Experiences in which a patient's expectations are not only met but exceeded, creating a sense of delight and excitement. |  |

For the quantitative data, descriptive statistical analyses were then performed, including the calculation of means and standard deviations for each stage of the hospitalization process, providing objective measures of patient satisfaction. This quantitative analysis offered a detailed understanding of patient experiences, identifying specific stages where patient satisfaction was either highest or lowest.

Qualitative information in the PJM was divided into two key aspects. First, it distinguished the factors that enhance the patient’s experience from those that detract from it; and second, it categorized the hospital’s strengths and areas for improvement. Additionally, quantitative results were presented as mean values, providing a clear, objective measure of patient satisfaction. Preliminary PJM, incorporating both qualitative themes and quantitative results, were presented to participants and key stakeholders for further validation and discussion.

## 3. Results

### 3.1. Stages of the Patient Journey

The hospitalization process was structured into five sequential stages: (1) Start of Hospitalization (emergency waiting, scheduled admission wait, ward admission), (2) Communal Living (nocturnal rest, awakening, hygiene/shower routines, meals), (3) Medical Process (medication administration, medical visits, non-pharmacological treatments), (4) Morning Activity (afternoon activities, door opening, therapeutic walks, family visits), and (5) End of Hospitalization (discharge process). These stages were validated through focus group feedback and expert panel consensus, forming the framework for analyzing patient experiences.

### 3.2. Qualitative Findings

The analysis of patient journey experiences identified a combination of positive elements and areas needing improvement throughout the hospitalization process. Many patients appreciated the attentive and compassionate care from staff, as reflected in comments such as, “They were always checking on me,” and, “They made me feel at ease right away.” Timely and clear communication before scheduled admissions was also valued: “They informed me in advance about my admission date,” and, “Staff were always kind when explaining things.” Conversely, prolonged waiting times and uncertainty, especially during admission from the emergency department, negatively affected patients: “I waited for hours without knowing what to expect,” and, “No one explained where I would be admitted.” Restrictions on personal belongings and strict security measures were often described as excessive: “I would have liked to have my phone,” and, “The security measures felt too strict.” Some patients reported feeling disoriented due to insufficient information about daily routines and unit rules: “I was confused about what I could do and the daily schedules,” and, “I didn’t know if I could wear my own clothes or not.”

Other frequently mentioned challenges included limited family visitation, discomfort related to room sharing and lack of privacy, and inconsistent information from medical and nursing teams. Despite these issues, patients consistently valued the warmth of staff, the cleanliness and modernity of facilities, and the availability of therapeutic activities. Overall, patients highlighted the need for clearer information, more flexible rules, and enhanced emotional support to further improve the hospital experience. In the PJM, these results were represented in two sections: first, patient-related issues (Figure 1), and second, hospital matters (Figure 2).

**Figure 1.**
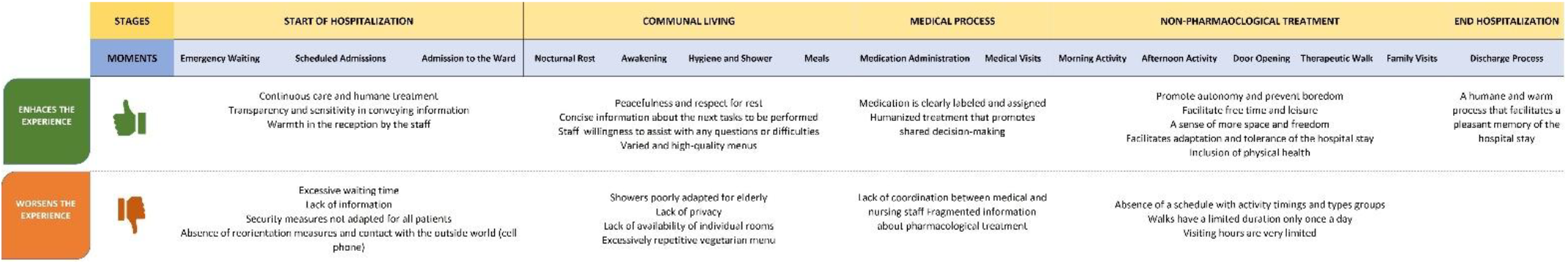
Patient-related issues presented in the PJM.

**Figure 2.**
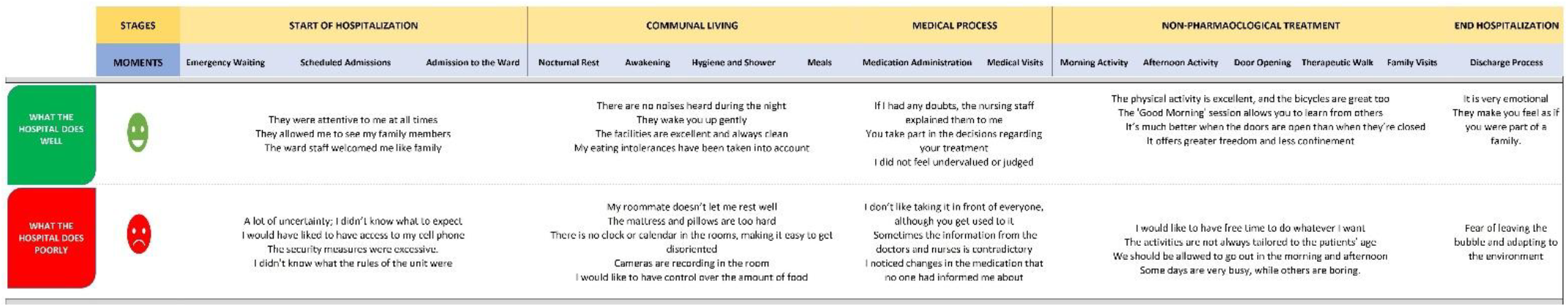
Hospital matters represented in the PJM.

Critical touchpoints were identified and categorized as: (1) Moments of Truth (ward admission, medical visits, door opening), where trust and collaboration shaped perceptions; (2) Moments of Pain (emergency waiting, hygiene routines, discharge anxiety), marked by frustration and systemic gaps; and (3) Wow Moments (medical visits, therapeutic walks), where patient expectations were exceeded through shared decision-making and autonomy promotion. They are represented in Figure 3.

**Figure 3.**
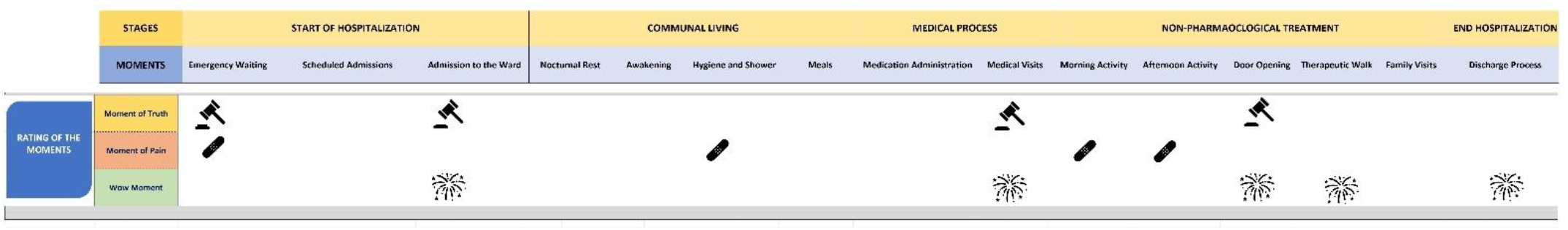
Critical touchpoints as part of the PJM.

### 3.3. Quantitative Outcomes

Mean satisfaction scores (1–6 Likert scale) varied significantly: the lowest interaction scores occurred during emergency waiting (3.8 ± 1.2) and afternoon activities (4.0 ± 0.9), while the highest scores were linked to medical visits (interaction: 5.7 ± 0.6; empathy: 5.8 ± 0.4) and discharge processes (empathy: 5.4 ± 0.7). Medication administration scored moderately (4.6 ± 1.1), reflecting discomfort with public routines, whereas communal living aspects like meals (4.8 ± 0.8) and nocturnal rest (5.3 ± 0.7 for empathy) received favorable ratings. It is also printed in the PJM, as shown in Figure 4.

**Figure 4.**
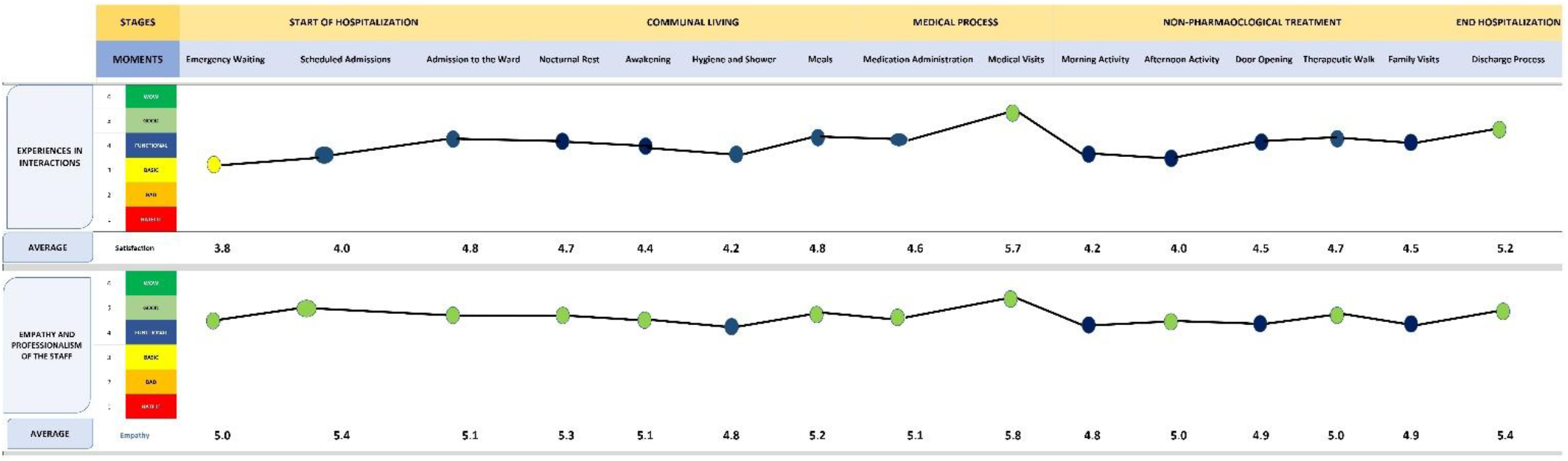
Quantitative results presented in the PJM.

### 3.4. Patient Journey Map

The final PJM, incorporating the study results, is presented in Figure 5. From top to bottom, it outlines, for each moment, factors that “Enhance” or “Worsen” the experience as well as the strengths and weaknesses of the hospital’s performance. It also provides the obtained ratings for Interaction and Empathy/Professionalism, helping to identify both strengths and areas needing improvement throughout the hospitalization process. In the bottom section, the key moments are represented using the symbols shown previously.

**Figure 5.**
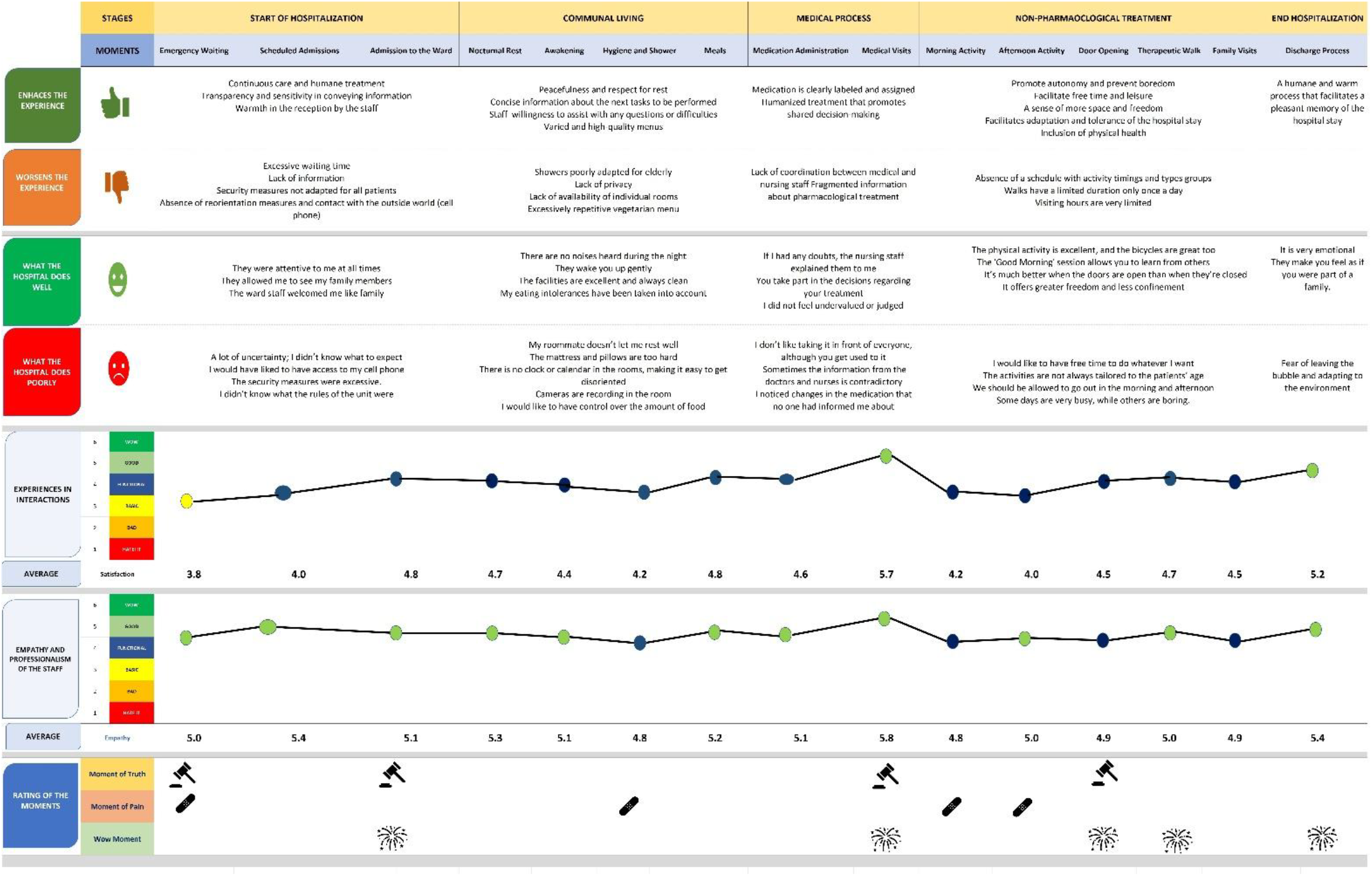
Final PJM.

## 4. Discussion

This study provides a comprehensive exploration of patient experiences within an open-door psychiatric unit, integrating qualitative narratives with quantitative data to elucidate critical aspects of the patient’s journey. By synthesizing both perspectives, the findings highlight key stages requiring improvement, particularly in admission processes, daily interactions, and therapeutic interventions, while underscoring the benefits of open-door policies in enhancing patient autonomy and satisfaction.

Qualitative feedback reinforces the quantitative trend, with patients frequently citing long waiting times and inadequate communication as significant stressors. Comments such as “I didn’t know what to expect” and “I waited for hours without updates” reflect these concerns and are consistent with the literature emphasizing the role of structured communication in reducing patient anxiety (Philpot et al., 2019; Murry, Al-Khatib and Witry, 2021). Implementing evidence-based communication protocols, such as routine updates and clear explanations of admission procedures, could mitigate these issues. Arias et al. further suggest leveraging technology, such as real-time status boards or mobile notifications, to enhance transparency during this critical phase. (Arias et al., 2020)

Patient satisfaction during medical visits and medication administration varied, with higher scores associated with shared decision-making and lower scores for medication routines. These findings are supported by prior research highlighting the importance of patient involvement in decision-making to foster engagement and satisfaction (Bakolis, Gupta and Wykes, 2022; Coelho et al., 2022). Comments such as “I felt heard and valued” illustrate the positive impact of empowering patients in their care. Conversely, dissatisfaction with medication routines often stemmed from issues of privacy and rigid scheduling, as reflected in statements like “I didn’t like taking it in front of everyone.” These concerns align with the literature documenting how perceived intrusions on dignity can negatively impact therapeutic experiences (Butterworth, Wood and Rowe, 2022; Kim and Nam, 2024). Strategies to address these issues include offering private spaces for medication administration and adopting more flexible schedules, which have been shown to improve patient satisfaction (Elgendy et al., 2023).

Therapeutic activities and the open-door model received some of the highest satisfaction scores. Patients frequently highlighted the benefits of autonomy and structured engagement, with comments such as “The freedom to walk outside made a big difference” and “Activities gave structure to my day.” These findings corroborate existing evidence that open-door policies enhance emotional well-being and reduce the psychological burden of hospitalization(Schneeberger et al., 2017; Hochstrasser et al., 2018). However, inconsistencies in the scheduling and communication of activities were reported, as illustrated by comments like “We don’t always know what’s planned for the day.” Practical solutions, such as digital or physical boards updated daily with activity schedules, have been suggested in the literature to improve transparency and predictability (Arias et al., 2020). Verbal reminders or mobile alerts could further ensure that patients are informed of any changes, thereby enhancing their engagement.

Discharge processes yielded the highest satisfaction scores, with patients emphasizing the role of empathetic interactions and clear guidance. Comments such as “It felt like leaving a family” and “Post-discharge instructions were very clear” reflect the effectiveness of structured discharge planning. These findings align with best practices outlined in frameworks such as the Transitional Care Model, which highlight the importance of tailored discharge plans and follow-up protocols to reduce post-discharge anxiety (Murry, Al-Khatib and Witry, 2021; Guilcher et al., 2024). Nonetheless, qualitative data revealed underlying concerns about adapting to external environments, echoing literature on post-discharge anxiety (Schouten et al.,2021). Strengthening follow-up systems, such as phone check-ins or digital support tools, could provide patients with additional reassurance and continuity of care.

Quantitative data indicate that patient satisfaction before admission was notably lower compared to other hospitalization stages. These findings align with prior studies demonstrating that waiting for admission in psychiatric settings, particularly delays for patients awaiting transfer from the emergency department, are often accompanied by heightened anxiety and stress due to uncertainty and a perceived lack of control (Georgieva, Mulder and Wierdsma, 2012; Wand et al., 2020; Wolff et al., 2023; Indregard et al., 2024). However, satisfaction scores during admission were consistent with those reported in other open-door units (Hochstrasser et al., 2018; Krückl et al., 2023).

This study has several strengths, including a comprehensive methodology that integrates qualitative and quantitative approaches, providing a robust framework for capturing both overarching trends and nuanced patient experiences. Additionally, its patient-centered focus aligns with global initiatives to humanize mental healthcare and improve therapeutic environments. However, certain limitations must be acknowledged. The generalizability of findings may be restricted due to the single-site design, highlighting the need for future research to replicate this methodology across diverse healthcare settings. Furthermore, the study is subject to potential response bias, as self-reported data may be influenced by social desirability or patient perceptions of staff. Triangulating patient-reported outcomes with observational or third-party evaluations could enhance the validity of the findings.

## 5. Conclusions

This study makes a significant contribution to the literature on patient experiences in open-door psychiatric units, integrating qualitative and quantitative findings to provide a holistic understanding of the challenges and benefits associated with this care model. By analyzing key aspects of the hospitalization process—including admission, daily interactions with staff, participation in therapeutic activities, and discharge transitions—the findings not only corroborate existing trends in the literature but also offer new insights into areas for improvement in psychiatric inpatient care. Future research should validate these findings in diverse settings and incorporate multi-stakeholder perspectives to further refine and optimize care delivery.

## Acknowledgments

The authors thank all participating volunteers for their contribution.

## Data Availability Statement

The dataset generated for this study is available upon request from the corresponding author.

## Notes

Funding: This work was supported by a grant from the JMC Legacy Research Fund of Germans Trias i Pujol University Hospital. It was also supported by the Fundació Llegat Roca i Pi. The sponsors had no role in the study design, data collection and analysis, interpretation of results, the preparation of the manuscript, the decision to submit the manuscript for publication, and the writing of the manuscript.

Conflicts of interest: The authors declare that the research was conducted in the absence of any commercial or financial relationships that could be construed as a potential conflict of interest.

### Competing Interest Statement

The authors have declared no competing interest.

### Funding Statement

This work was supported by a grant from the JMC Legacy Research Fund of Germans Trias i Pujol University Hospital. It was also supported by the Fundacio Llegat Roca i Pi. The sponsors had no role in the study design, data collection and analysis, interpretation of results, the preparation of the manuscript, the decision to submit the manuscript for publication, and the writing of the manuscript.

### Author Declarations

Ethics committee of Hospital Universitari Germans Trias i Pujol gave ethical approval for this work

